# Arterial spin labeling as a promising alternative to FDG-PET for clinical diagnosis of patients with disorders of consciousness

**DOI:** 10.1101/2024.08.07.24311648

**Authors:** Junrong Han, Niall W. Duncan, Likai Huang, Chia Lee, Che-Ming Yang, Yen-Chien Wu, David Yen-Ting Chen, Timothy J. Lane

## Abstract

**Objective:** To evaluate the potential of arterial spin labeling (ASL) as an alternative to FDG-PET in the diagnosis of disorders of consciousness (DOC), we conducted a comparative study of the two modalities.

**Methods:** A total of 36 DOC patients (11 female; mean age = 49.67 ± 14.54 years) and 17 healthy control (HC) participants (9 female; mean age = 31.9 ± 9.6 years) underwent both FDG-PET scans that measure metabolism via glucose uptake and ASL scans that measure cerebral blood flow (CBF). CBF and metabolism in DOC and HC were compared, globally and for seven functional networks. Comparability of the two modalities was estimated using Spearman partial correlation for both global and network levels. Furthermore, a support vector machine (SVM) algorithm was used to train two classifiers to distinguish DOC patients from HC with normalized CBF or metabolism values from the seven networks. Performance of these classifiers was first evaluated through leave-one-subject-out (LOSO) cross-validation within their respective modalities. Subsequently, cross-modal validation was conducted: testing the ASL-trained classifier with PET data (ASL-to-PET validation) and vice versa (PET-to-ASL validation). Performance of each classifier was assessed using receiver operating characteristic (ROC) analysis, with area under the curve (AUC) as the metric.

**Results:** Both modalities showed agreement in decreased CBF and metabolism in DOC patients compared to HC, at both global and network levels. The global brain and most networks showed significant positive partial correlation between CBF and metabolism. SVM classifiers, utilizing activity from seven networks as features, performed well in the both LOSO cross-validation (ASL-trained classifier: accuracy = 83.02%, AUC = 0.95; PET-trained classifier: accuracy = 98.12%, AUC = 0.98) and cross-modal validation (ASL-to-PET validation: accuracy = 84.90%, AUC =0.92; PET-to-ASL validation: accuracy = 73.58%, AUC = 0.93).

**Conclusion:** Our results demonstrate that ASL provides information comparable to FDG-PET for DOC patients; moreover, classifiers trained on ASL data perform comparably well to those trained on FDG-PET data. Therefore, ASL could be a valuable alternative to FDG-PET in the clinical diagnosis of DOC, especially in light of its advantages: ease of acquisition, avoidance of radiation exposure, brevity of scanning time, and lower-cost.

## INTRODUCTION

Disorders of consciousness (DOC) are characterized by a reduction in or absence of awareness of oneself and the surrounding environment, typically caused by traumatic brain injury, cerebrovascular accident, hypoxia or anoxia^1^. The Coma Recovery Scale—Revised (CRS-R)^2^ can be used to classify these patients based on their behavioral features into either unresponsive wakefulness syndrome (UWS) patients^3^, who suffer complete loss of consciousness, or minimally consciousness state (MCS) patients^4^, who exhibit some, but often unstable, behavioral indications of awareness. Accurate diagnosis of DOC is essential to determining appropriate treatment and prognosis, as well as improving quality of life for patients. Despite the promising performance of 18F-fluorodeoxyglucose positron emission tomography (FDG-PET) in diagnosing DOC patients by measuring cerebral glucose metabolism^5–11^, its high cost, the need to administer a radioactive tracer, the speed of tracer decay, and other factors combine to limit its clinical application.

Use of arterial spin labeling (ASL) to quantify cerebral blood flow (CBF)^12,13^ has shown promise as a non-invasive alternative to FDG-PET in measuring neuronal activity in healthy individuals^14–16^. ASL possesses several advantages, making it a viable option for clinical usage: it is nonradioactive and can be carried out at relatively low cost, with a brief acquisition time, on MRI scanners that are widely available, and it does not require fasting. Studies have also shown that ASL can identify brain abnormalities that are also identified by FDG-PET; these include markers of neurodegenerative diseases such as Alzheimer’s disease and frontal-temporal dementia^17–20^. Of direct relevance to our investigation, several brain regions with impaired CBF in DOC patients, such as the prefrontal and posterior cortices, comport with FDG-PET findings for DOC patients^5–11,21,22^. However, direct evidence for the conjecture that ASL can substitute for FDG-PET in the diagnosis of DOC patients is insufficient. Accordingly, this study investigates the DOC diagnostic comparability of these two modalities.

To achieve this aim, we collected ASL and FDG-PET data from both DOC patients and healthy controls (HC). At the global level, we compared average CBF to an FDG-PET metabolic index between DOC and HC, and evaluated their partial correlation across all participants. Additionally, we performed the same comparisons for seven functional networks^23^. To compare the clinical performance of the two modalities, we used normalized metrics from the seven networks as features to train support vector machine (SVM) classifiers. Leave-one-subject-out (LOSO) cross-validation and cross-modal validation were used to evaluate classifier performance. Finally, receiver operating characteristic (ROC) analysis was conducted to estimate the classification performance for each validation method. Fig.1 illustrates the overall processing procedure.

**Fig.1.**
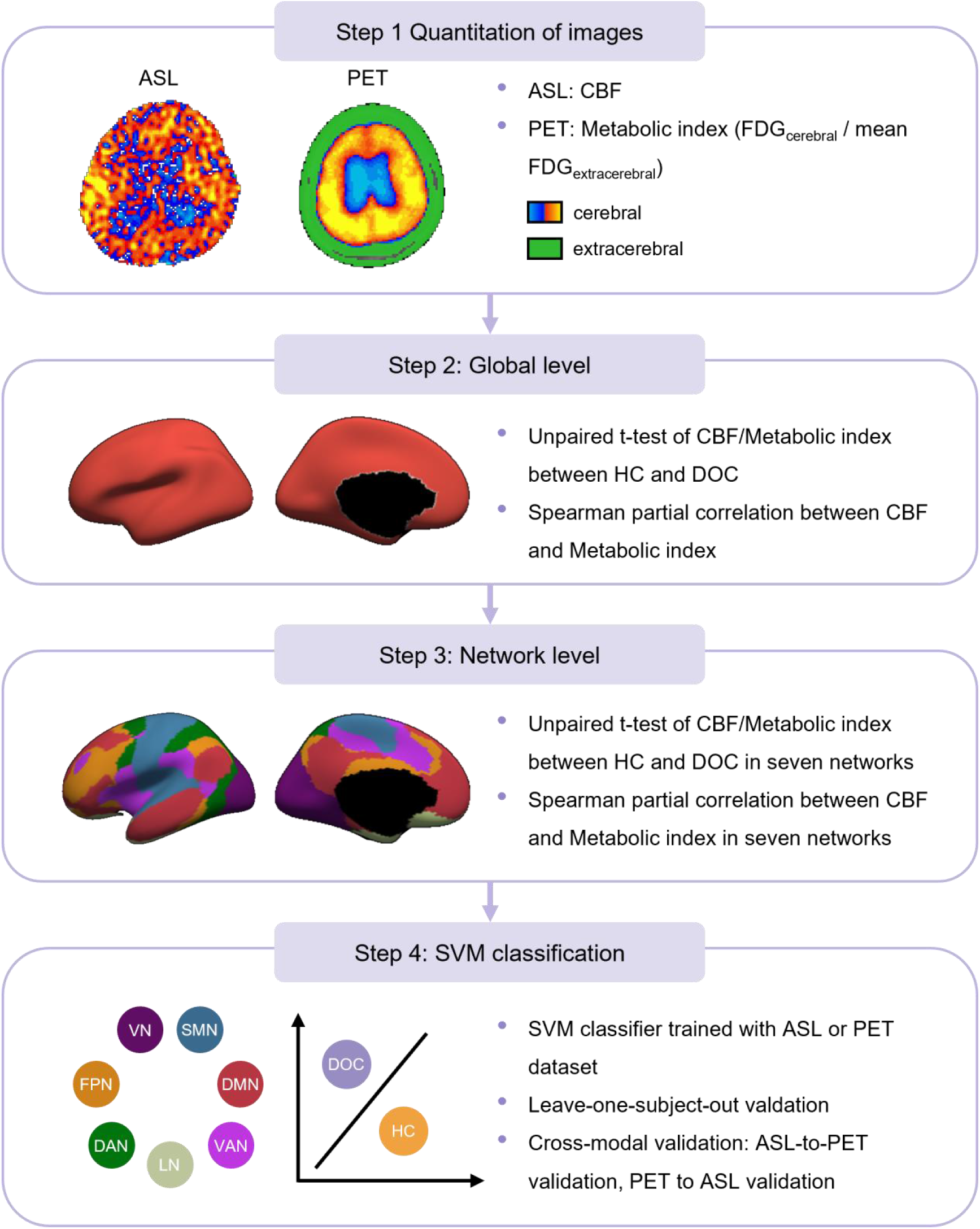
Flowchart of data processing procedure. Step 1 involves quantifying the images in both ASL and FDG-PET data. Step 2 involves comparing the cerebral blood flow (CBF) or metabolic index between HC and DOC groups at the global level, and performing Spearman’s partial correlation between the two modalities at the global level. Step 3 involves comparing the CBF or metabolic index between HC and DOC at the network level, and performing Spearman’s partial correlation between the two modalities at the network level. Step 4 involves model training and cross-validation for SVM classifiers. DMN = default mode network, FPN = frontoparietal network, LN = limbic network, VAN = ventral attention network, DAN = dorsal attention network, SMN = somatomotor network, VN = visual network.

## METHODS

### Patient Recruitment

Eighty-one DOC patients were recruited. Of these, 36 completed the required combination of MRI and FDG-PET scanning and had usable data (11 female; mean age = 49.7 ± 14.5 years). Of these 36 patients, 33 were classified as UWS and three as MCS. Clinical assessment was conducted by qualified physicians and others specifically trained in use of the CRS-R instrument. Assessments were repeated multiple times at different times of day and under varied circumstances, with patients in a sitting posture where possible. The etiologies of DOC included traumatic brain injury, cerebrovascular accident, hypoxia, anoxia, and carbon monoxide poisoning. Further information regarding patient diagnosis can be found in Table 1. Additionally, 17 healthy individuals (9 female; mean age = 31.9 ± 9.6 years) were recruited as a control group. None of the control participants had a history of neurological or psychiatric disorders, nor were they taking medication at the time of the scan.

**Table 1.**
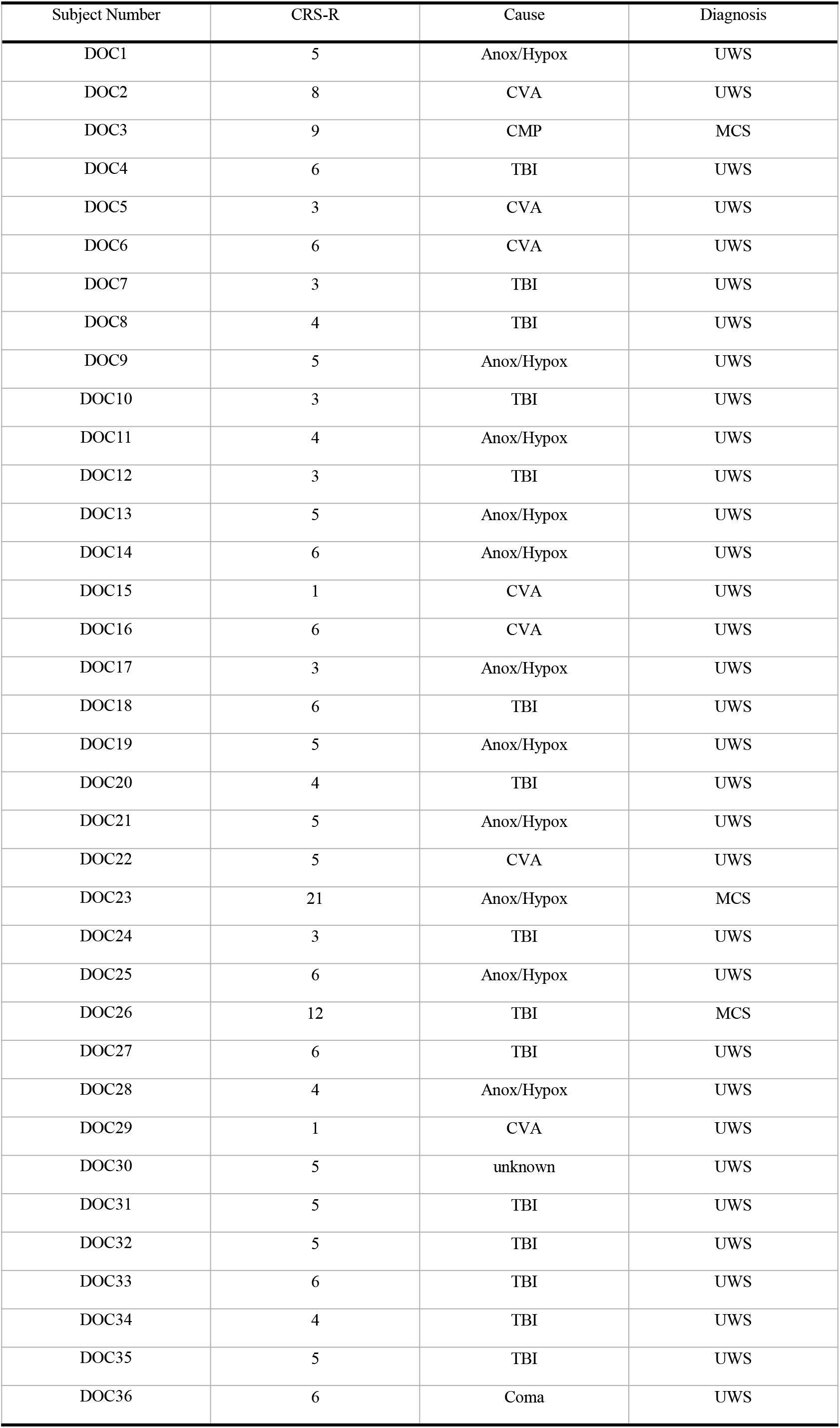
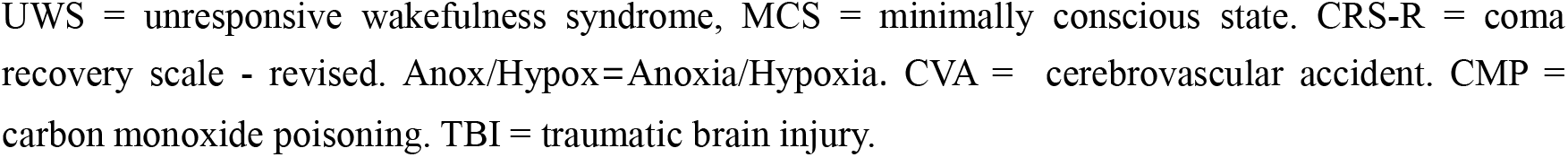
Clinical information for patients with disorders of consciousness.

| Subject Number | CRS-R | Cause | Diagnosis |
| --- | --- | --- | --- |
| DOC1 | 5 | Anox/Hypox | UWS |
| DOC2 | 8 | CVA | UWS |
| DOC3 | 9 | CMP | MCS |
| DOC4 | 6 | TBI | UWS |
| DOC5 | 3 | CVA | UWS |
| DOC6 | 6 | CVA | UWS |
| DOC7 | 3 | TBI | UWS |
| DOC8 | 4 | TBI | UWS |
| DOC9 | 5 | Anox/Hypox | UWS |
| DOC10 | 3 | TBI | UWS |
| DOC11 | 4 | Anox/Hypox | UWS |
| DOC12 | 3 | TBI | UWS |
| DOC13 | 5 | Anox/Hypox | UWS |
| DOC14 | 6 | Anox/Hypox | UWS |
| DOC15 | 1 | CVA | UWS |
| DOC16 | 6 | CVA | UWS |
| DOC17 | 3 | Anox/Hypox | UWS |
| DOC18 | 6 | TBI | UWS |
| DOC19 | 5 | Anox/Hypox | UWS |
| DOC20 | 4 | TBI | UWS |
| DOC21 | 5 | Anox/Hypox | UWS |
| DOC22 | 5 | CVA | UWS |
| DOC23 | 21 | Anox/Hypox | MCS |
| DOC24 | 3 | TBI | UWS |
| DOC25 | 6 | Anox/Hypox | UWS |
| DOC26 | 12 | TBI | MCS |
| DOC27 | 6 | TBI | UWS |
| DOC28 | 4 | Anox/Hypox | UWS |
| DOC29 | 1 | CVA | UWS |
| DOC30 | 5 | unknown | UWS |
| DOC31 | 5 | TBI | UWS |
| DOC32 | 5 | TBI | UWS |
| DOC33 | 6 | TBI | UWS |
| DOC34 | 4 | TBI | UWS |
| DOC35 | 5 | TBI | UWS |
| DOC36 | 6 | Coma | UWS |

UWS = unresponsive wakefulness syndrome, MCS = minimally conscious state. CRS-R = coma recovery scale - revised. Anox/Hypox=Anoxia/Hypoxia. CVA = cerebrovascular accident. CMP = carbon monoxide poisoning. TBI = traumatic brain injury.

This study was approved by the TMU-Joint Institutional Review Board (N202204087 and N202109034). Informed, written consent was granted by HCs and by family members or legal guardians of DOC patients. DOC patients were recruited from all parts of Taiwan, under the aegis of Taipei Medical University—Shuang Ho Hospital’s Brain and Consciousness Research Centre.

### Data acquisition

FDG-PET data were acquired on a GE Discovery ST PET-CT scanner. HCs were asked to fast for eight hours prior to the scan session. [^18^F]-fluorodeoxyglucose (mean dose = 11.8 mCi±1.2 SD) was administered intravenously, after which participants rested in a darkened room for 40 min. They were asked to lie with their eyes closed during this time. A 20-min scan was then conducted with the eyes closed. The same general protocol was used for DOC patients. DOC scans were scheduled for early morning to approximate an eight hour fast in a manner that did not interfere with their normal care.

ASL data were acquired on a GE MR750 3 Telsa scanner using a standard 8-channel head coil. A high-resolution T1-weighted anatomical image was acquired first. Following this, CBF images were acquired using a 3D pCASL ASL sequence with a fast spin echo acquisition for vessel suppression (TR = 5327 ms; TE = 10.5 ms; FoV = 220 mm; slice thickness = 4 mm; slice gap = 0 mm; 38 slices; NEX = 4; labeling duration = 1500 ms; post-labeling delay = 1525 ms; scan duration = 6.63 min). Participants were instructed to lie still with eyes closed during the scan. Scans were conducted at the same time of day for HCs but varied for DOC patients, in accordance with clinical and logistic factors. ASL and FDG-PET data from the HC group were included in a previously published analysis^24^.

### Definition of regions of interest

Identifying small regions within the impaired brains of DOC patients presents significant challenges. To address this, we targeted a set of specific regions of interest (ROIs), namely: the total cortical gray matter; seven predefined networks^23^; and 100 cortical areas allocated to these networks^25^. We utilized FSL tools (https://fsl.fmrib.ox.ac.uk/fsl/fslwiki/) to create individual ROIs for each participant by wrapping the ROIs from MNI standard space into individual space. The processing procedures were as follows: First, we extracted the ROIs from the MNI standard brain to obtain the ROI templates. Next, we registered the FDG and CBF images of each participant to the individual structural images to obtain the spatial transformation matrix of FDG-to-structure and CBF-to-structure. We then normalized the individual structural image to MNI standard space to obtain a structure-to-MNI transformation matrix. The transformation matrices were then inverted and combined to produce MNI-to-FDG and MNI-to-CBF transforms. These were then applied to align the different ROI templates to the individual FDG and CBF images.

To diminish the influence from cerebrospinal fluid (CSF) and seriously damaged brain tissue on our results, we excluded these regions from the aligned ROIs. To achieve this, we first utilized the MNI-to-FDG or MNI-to-CBF transformations to warp the MNI standard CSF template into individual image spaces. Areas of seriously damaged brain tissue were then manually identified in individual structural images and warped to the individual FDG or CBF spaces using the structure-to-FDG or structure-to-CBF transformations, respectively. Finally, areas of CSF and damaged brain tissue were removed from the ROIs in individual space.

### Image quantification and data extraction

To reduce the influence of brain injury on the FDG-PET images in DOC patients, we adopted a pseudo-quantitative method that involved normalizing cerebral FDG values to the intensity of the extra-cerebral tissue^9^. Specifically, cerebral FDG-PET values were divided by those averaged across extra-cerebral tissue, where the intensity ranged from the 5th to 95th percentile, to generate a metabolic index image for each participant. The metabolic index within each aligned ROI was then averaged. For ASL data extraction, the CBF within each aligned ROI was averaged to obtain CBF for each participant.

### Global- and network-level modality comparison

To investigate the relationship between CBF and FDG metabolism, we contrasted these two modalities between participant groups (HC and DOC) and calculated correlations at both global and network levels. At the global level, unpaired t-tests were performed to compare grey matter CBF and the metabolic index between HC and DOC patients. Spearman partial correlation was then calculated between grey matter CBF and metabolic index across all subjects A group factor (viz. DOC and HC) was included as a covariate in these correlations so that outcomes were not driven by the overall between-group differences in CBF and metabolic index. At the network level, the same unpaired t-test and Spearman partial correlation analyses were conducted for each network. False discovery rate (FDR) correction was performed for a total of eight unpaired t-tests and for eight partial correlations.

### Classification analysis

To explore the potential of ASL as a reliable alternative to FDG-PET in clinical diagnosis of DOC, we constructed two SVM classifiers with the ASL and PET data and evaluated their classification performance. Each classifier comprised the CBF or metabolic index values from the seven networks, giving seven features per individual in each classifier. Since CBF measured by ASL and metabolism measured by PET have different reference baselines, the SVM classifier trained on ASL data or PET data could not be directly tested with data from the other modality, making it difficult to validate ASL as a replacement for PET. Therefore, we normalized the features for each participant by subtracting the mean and dividing by standard deviation across all features. This normalization enabled the training of the ASL-based classifier with adjusted CBF features and the PET-based classifier with adjusted metabolic features. The classifier was initially assessed via LOSO cross-validation within their respective modalities. Subsequent cross-modal validation involved testing the ASL-trained classifier with normalized PET data (ASL-to-PET validation) and the PET-trained classifier with normalized ASL data (PET-to-ASL validation). To further evaluate robustness, classifiers were also trained using another set of 100 features derived from 100 cortical areas for both CBF (ASL) and metabolic index (PET). These classifiers underwent the same LOSO cross-validation and cross-modal validation procedures.

Performance of all SVM classifiers was analyzed quantitatively using confusion matrices and ROC analysis to assess their classification accuracy and discriminative ability between DOC patients and HCs. Specifically, classification accuracy was calculated by adding true positive rates and true negative rates in a confusion matrix; the area-under-the-curve (AUC) was computed by ROC analysis to estimate classification performance.

### Data Availability

The source data for figures will be publicly available at Zenodo as of the date of publication.

## RESULTS

### Global-level modality comparison

Grey matter CBF and metabolic index values for each group are presented in Table 2. An unpaired t-test showed a decrease in both the metabolic index and CBF for global grey matter in DOC patients, compared to HC (t_PET_ = 14.88, p < 0.001, FDR corrected; t_ASL_ = 12.55, p < 0.001, FDR corrected) (Fig.2A). While controlling for overall group differences, Spearman partial correlation showed a positive correlation of 0.41 between the mean CBF and mean metabolic index within grey matter across all participants (two-tailed test, p < 0.05, FDR corrected) (Fig.2B).

**Table 2.** Mean CBF and metabolic index of ROIs in different group.

| ROI | CBF (mean ± S.D) | | Metabolic index (mean ± S.D) | |
| --- | --- | --- | --- | --- |
|  | HC | DOC | HC | DOC |
| Global gray matter | 52.05 ± 10.25 | 17.70 ± 8.83 | 9.07 ± 1.25 | 4.23 ± 1.03 |
| VN | 45.10 ± 9.68 | 14.56 ± 8.00 | 9.11 ± 1.37 | 4.11 ± 1.03 |
| SMN | 50.45 ± 9.90 | 18.87 ± 9.01 | 8.70 ± 1.31 | 4.47 ± 1.16 |
| DAN | 48.06 ± 11.16 | 15.02 ± 8.13 | 9.10 ± 1.40 | 4.27 ± 1.07 |
| VAN | 55.64 ± 10.32 | 19.64 ± 9.85 | 8.99 ± 1.27 | 4.27 ± 1.11 |
| LN | 48.76 ± 9.71 | 19.64 ± 9.21 | 7.39 ± 0.90 | 3.55 ± 0.93 |
| FPN | 58.15 ± 12.04 | 17.99 ± 9.73 | 9.68 ± 1.38 | 4.24 ± 1.19 |
| DMN | 56.70 ± 11.19 | 19.05 ± 9.52 | 9.15 ± 1.28 | 4.14 ± 1.05 |

**Fig.2.**
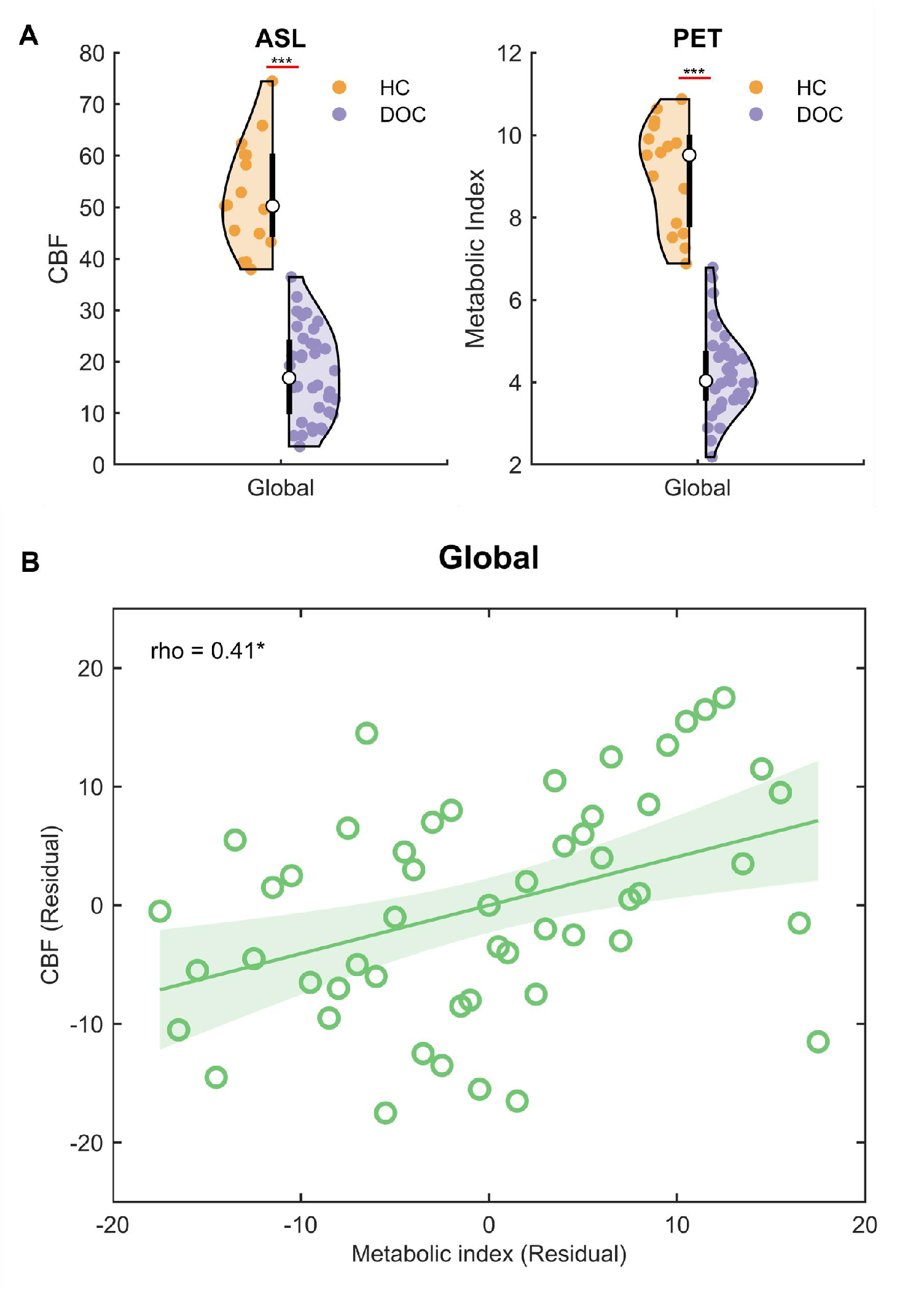
A) Unpaired t-test of CBF and metaboloc index between HC and DOC in global gray matter (FDR corrected, *** indicates p < 0.001). B) Spearman’s partial correlation between CBF and metabolic index across all participants in global gray matter (two-tailed test, FDR corrected, * indicates p < 0.05).

### Network-level modality comparison

CBF and metabolic index values are shown for each network in Table 2. Unpaired t-tests showed decreases in CBF and metabolic index in DOC patients, compared to HC, in all networks (PET: t_VN_ = 14.87, t_SMN_ = 11.89, t_DAN_ = 13.81, t_VAN_ = 13.79, t_LN_ = 14.14, t_FPN_ = 14.78, t_DMN_ = 15.10, p < 0.001, FDR corrected; ASL: t_VN_ = 12.11, t_SMN_ = 11.54, t_DAN_ = 12.22, t_VAN_ = 12.23, t_LN_ = 10.56, t_FPN_ = 12.98, t_DMN_ = 12.71, p < 0.001, FDR corrected) (Fig.3A,B).

**Fig.3.**
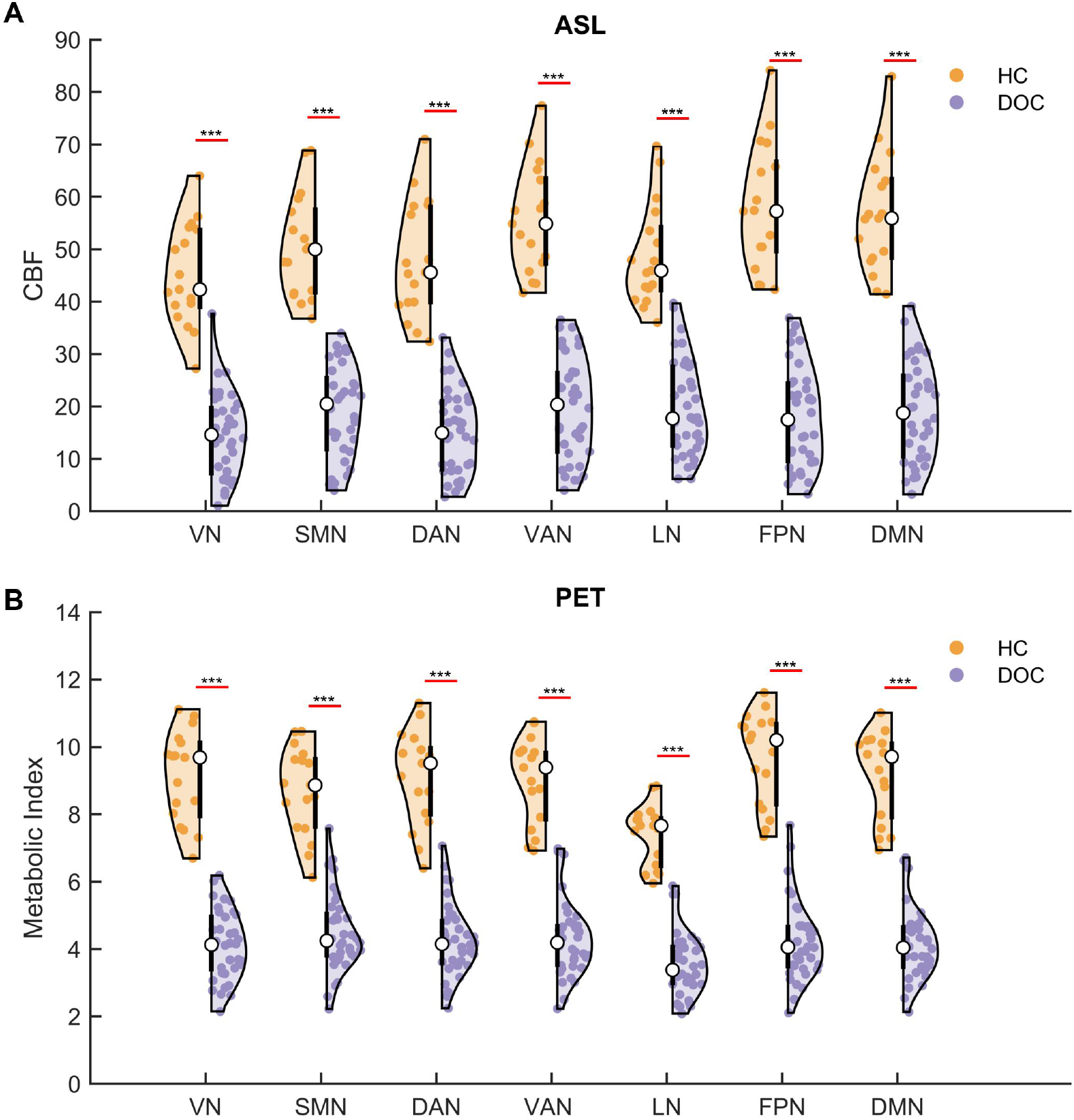
A) Unpaired t-test of CBF between HC and DOC in seven networks (FDR corrected, *** indicates p < 0.001). B) Unpaired t-test of metabolic index between HC and DOC in seven networks (FDR corrected, *** indicates p<0.001).

As shown in Fig.4, most networks showed positive partial correlations between CBF and metabolic index across all participants (rho_SMN_ = 0.36, rho_DAN_ = 0.35, rho_VAN_ = 0.42, rho_LN_ = 0.46, rho_FPN_ = 0.42, rho_DMN_ = 0.44; two-tailed tests, all p < 0.05, FDR corrected). The exception was the VN, where no correlation was found (two-tailed tests, p > 0.05, FDR corrected).

**Fig.4.**
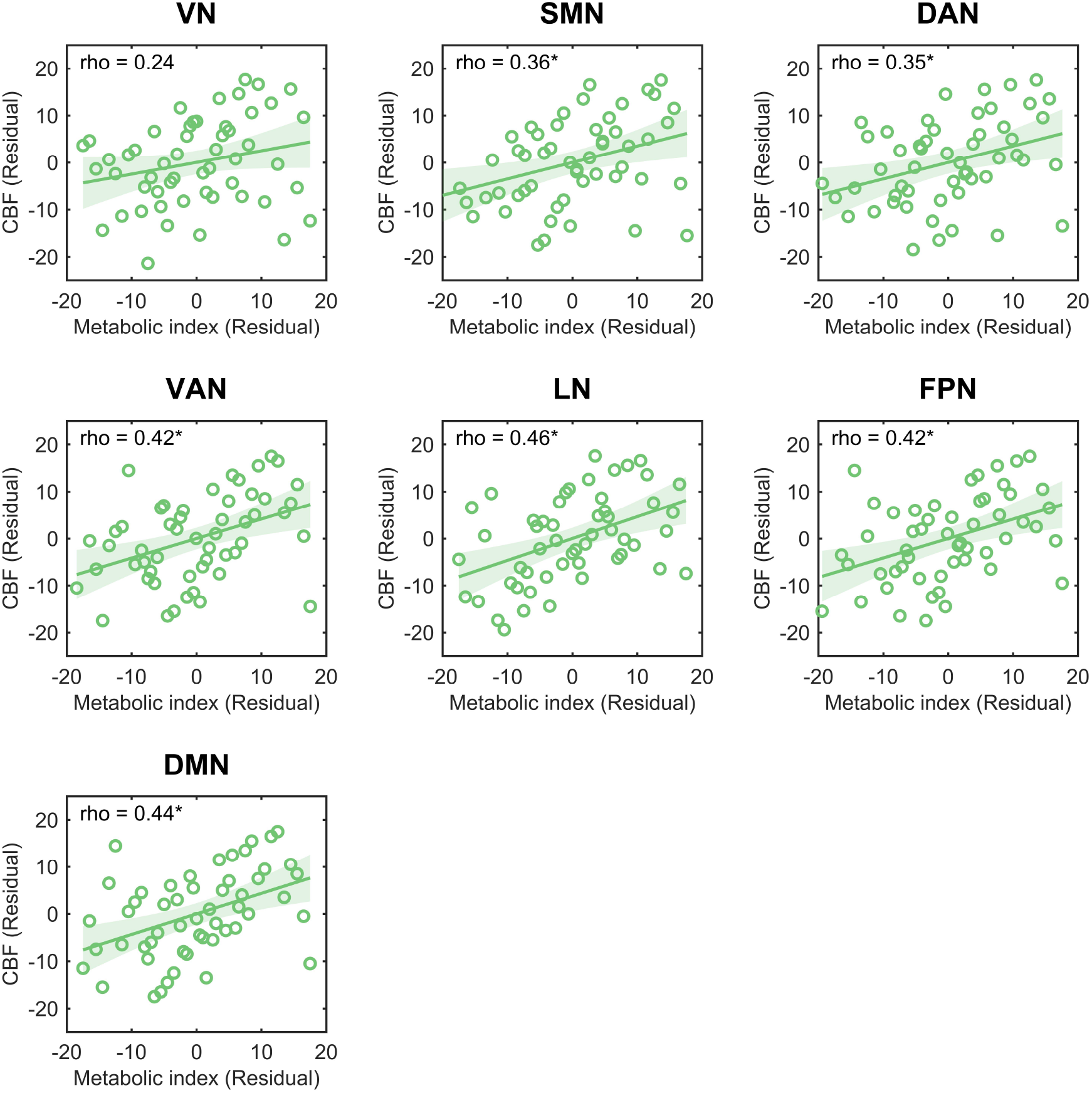
Spearman partial correlation between CBF and metabolic index across all participants in seven networks (two-tailed test, FDR corrected, * indicates p<0.05).

### DOC and HC classifier performance

Separate SVM classifiers were trained on seven network features from both the ASL and the FDG-PET data to discriminate between DOC patients and HC. Performance was tested using LOSO cross-validation and cross-modal validation. The ASL-trained classifier proved effective at discriminating between the two groups (accuracy = 83.02%; AUC = 0.95; Fig.5A); likewise the classifier trained on the PET data (accuracy = 98.12%; AUC = 0.98; Fig.5B). Notably, the ASL-trained classifier also performed well in differentiating between the groups using ASL-to-PET validation (accuracy = 84.90%; AUC = 0.92; Fig.5C), while the PET-trained classifier displayed robust discrimination in PET-to-ASL validation (accuracy = 73.58%; AUC = 0.93; Fig.5D). The distance from the decision boundary for each individual, reflecting the individual classification results for each cross-validation approach, is illustrated in the middle column of Fig.5A-D.

**Fig.5.**
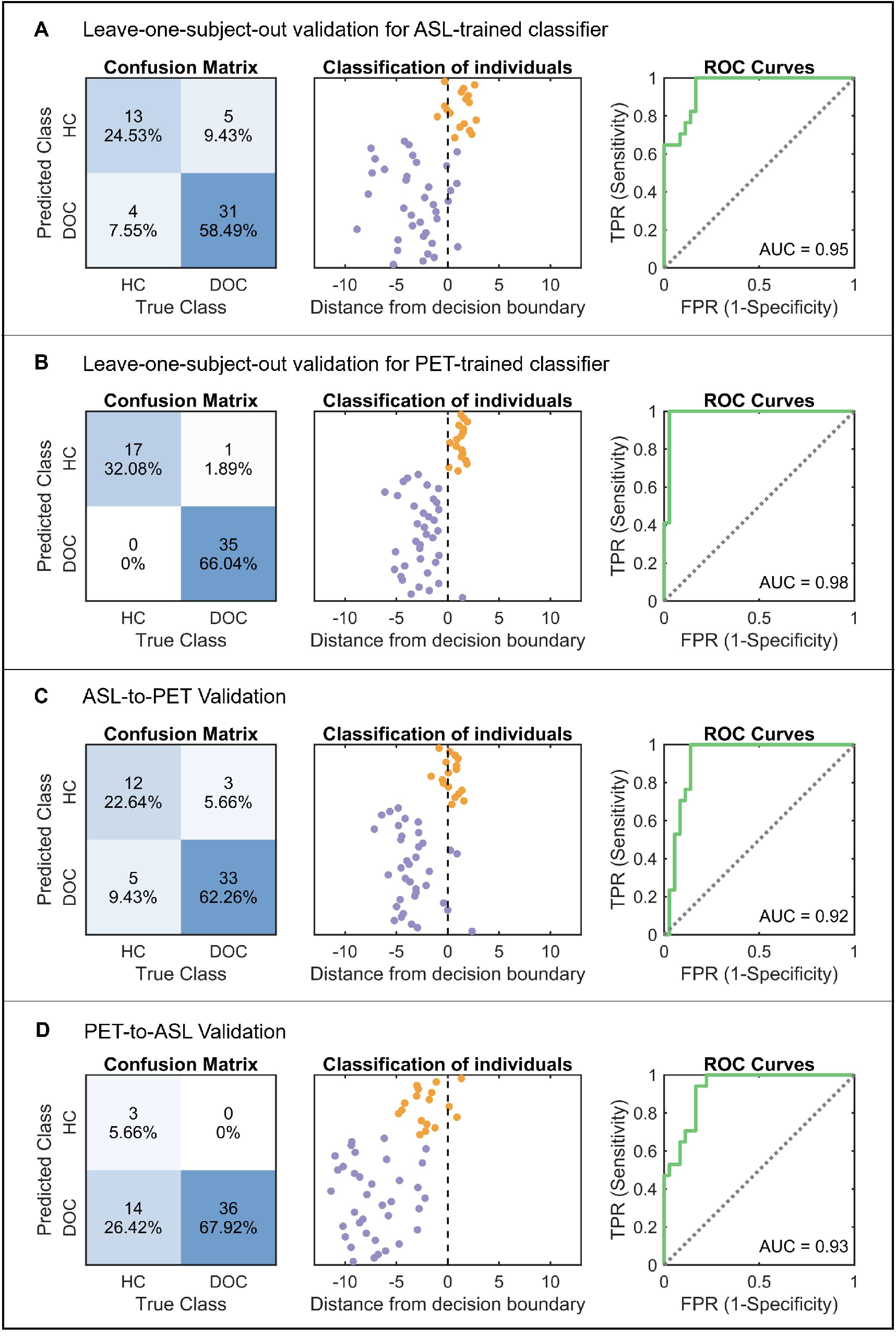
Performance of SVM classifiers trained with seven network features. A) The ASL-trained and B) PET-trained classifiers were validated using leave-one-subject-out cross-validation. C) The ASL-trained classifier was validated with the normalized metabolic index of seven networks in PET data. D) The PET-trained classifier was validated with the normalized CBF of seven networks in ASL data. Confusion matrices, individual classification result and ROC curve are displayed for each validation. ROC = receiver operating characteristic; AUC = area under curve. TPR = true positive rate; FPR = false positive rate.

To further explore classifier performance, we trained classifiers using data from each modality, extracting 100 features from 100 cortical regions, and conducted both LOSO cross-validation and cross-modal validation for each classifier. Both ASL-trained and PET-trained classifiers performed well when differentiating participant groups, as measured through LOSO cross-validation (ASL: accuracy = 98.12%, AUC = 1; PET: accuracy = 100%, AUC = 1) (Fig.6A-B). Moreover, both classifiers exhibited strong generalizability in cross-modal validation (ASL-to-PET: accuracy = 92.46%, AUC = 1; PET-to-ASL: accuracy = 77.35%, AUC = 0.99) (Fig.6C-D). These findings suggest that both classifiers evince robust performance in diagnosing DOC, and that an SVM classifier trained on one modality can provide classifications comparable to the other modality. Individual classification performance is shown in the middle column of Fig.6A-D.

**Fig.6.**
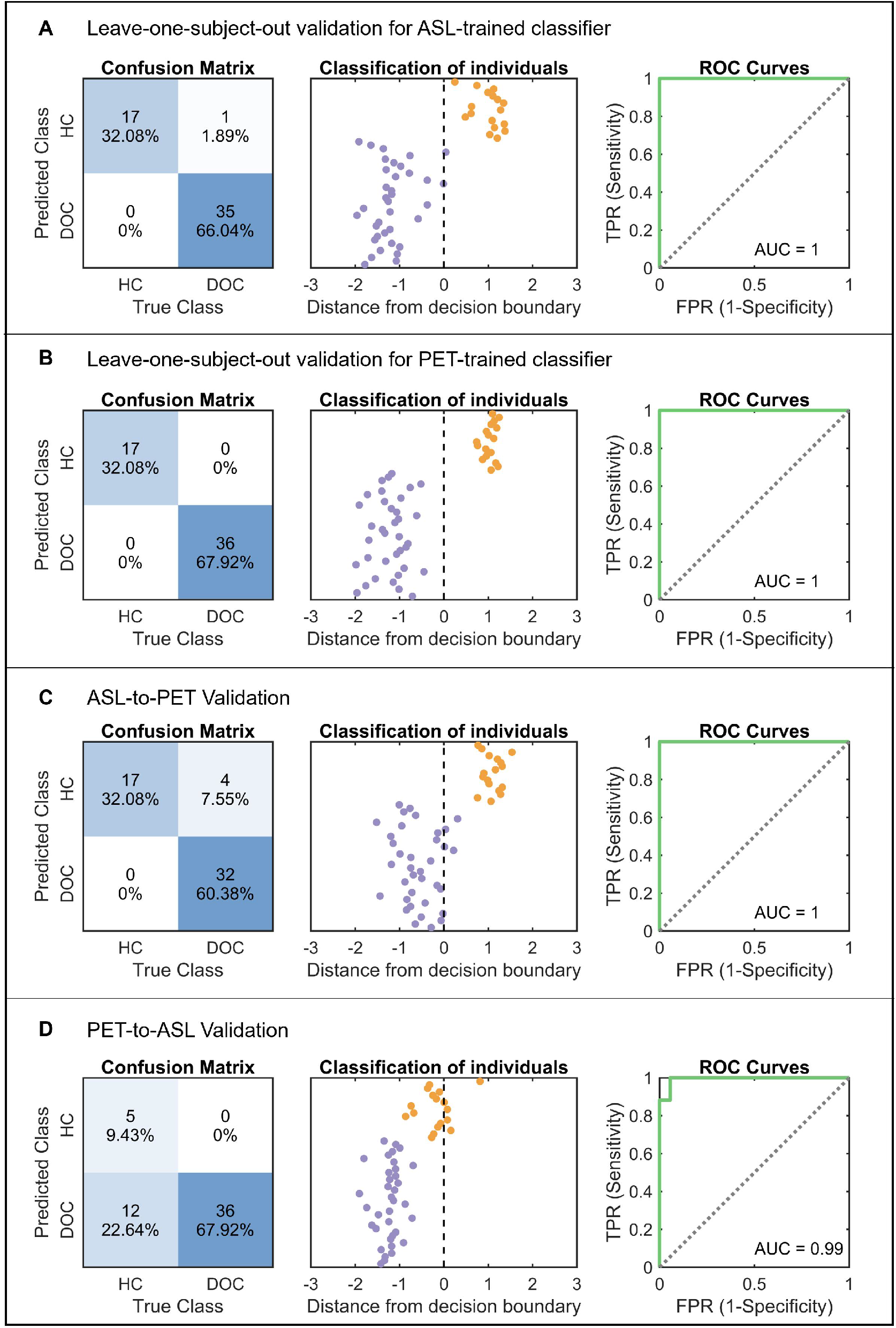
Performance of SVM classifiers trained with 100 features from 100 cortical regions. A) The ASL-trained and B) PET-trained classifiers were validated using leave-one-subject-out cross-validation. C) The ASL-trained classifier was validated with the normalized metabolic index from 100 regions in PET data. D) The PET-trained classifier was validated with the normalized CBF from 100 regions in ASL data. Confusion matrices, individual classification result and ROC curve are displayed for each validation. ROC = receiver operating characteristic; AUC = area under curve. TPR = true positive rate; FPR = false positive rate.

## DISCUSSION

The study reported here investigated whether MRI measures of CBF could be used as a valid proxy for metabolism in the diagnosis of DOC patients. Decreased CBF and metabolic indices in DOC patients compared to HCs were observed, both globally and for each of seven networks. Furthermore, positive correlations between CBF and metabolic indices were observed, both globally and in most functional networks. Importantly, SVM classifiers performed well when discriminating between DOC patients and HCs, as shown by the high accuracy rates and AUC values. The ASL-to-PET validation yielded an accuracy rate of 92.73% and an AUC of 0.94, while the PET-to-ASL validation yielded an accuracy rate of 87.27% and an AUC of 0.96.

Clinical application of FDG-PET for aiding in diagnosis of DOC patients has been well established. Previous studies comparing DOC patients to HCs have reported decreased metabolic activity globally as well as in the DMN, findings that are consistent with this investigation ^7–9,26,27^. However, the clinical utility of FDG-PET is limited, because it incurs several costs and risks: exposure to radiation, complexities pertaining to synthesis and administration of the radioactive FDG tracer, lengthy preparation (e.g. six to eight hours of fasting) and scanning times, and high cost. ASL, on the other hand, offers several advantages over FDG-PET: it is non-radioactive, non-invasive, more accessible, less costly, less time-consuming (approximately six minutes), and doesn’t require a preparatory fast. Furthermore, evidence suggests that CBF and metabolism are related, at the level of function^14–16,28^. Indeed, ASL and FDG-PET investigations of Alzheimer’s Disease have identified similar patterns of cerebral deficit^17–19,28,29^, and ASL has proven to be of clinical value in the assessment of several other brain disorders, such as stroke, post-traumatic stress disorder, and epilepsy^29^. To the best of our knowledge, the present study is the first to adduce evidence showing that ASL compares favorably to FDG-PET in the diagnosis of DOC. In a word, ASL could serve as a viable alternative to FDG-PET to aid in diagnosis of DOC.

This study also presents a novel use of the SVM algorithm for classification, when comparing measurements derived from two distinct modalities (ASL and FDG-PET in this case). Because CBF is measured by ASL, an MRI technique that involves magnetic labeling of water molecules^13^, while glucose metabolism is measured by FDG-PET which involves use of a radio-labelled tracer analogous to glucose, the two methods are not commensurable. Therefore, it is not possible to validate directly the SVM classifier trained on one modality dataset to a classifier trained on a distinct modality dataset. To address this problem and adduce more evidence for the reliability of ASL as a substitute for FDG-PET in clinical settings, we proceeded as follows: calculate the normalized activity of seven networks for datasets in both modalities; characterize the contribution rate of each network to the mean activity across all networks; and unify the baseline activity of all networks across both modalities. By proceeding in this way, the SVM classifier trained on the normalized network activity in ASL data could be validated by PET data, and in like manner the SVM classifier trained on PET data could be used to validate the ASL data. Results derived from using this method provide compelling evidence that ASL can substitute for FDG-PET in the diagnosis of DOC patients. What is more, we believe this method could be applied generally, to other investigations that involve classification performance of incommensurate modalities.

In previous ASL studies of DOC patients, reduced CBF was detected in several regions: the medial frontal and mid-frontal regions in MCS compared to HC^22^, as well as the putamen, anterior cingulate gyrus, and medial frontal regions in UWS compared to MCS^21^. However, little is known about the alteration patterns of CBF across multiple networks in DOC patients. In this study, we detected hypo-perfusion of cerebral blood flow in seven large-scale networks, reflecting an apparent global reduction of cortical activity that is consistent with the FDG-PET results. Our findings suggest a global reduction in CBF and metabolism in DOC patients, rather than reduction limited to a single region or network, such as the DMN, which has been postulated to be a neural substrate for consciousness as well as an indicator of the likelihood of recovery of a capacity for consciousness^30,31^.

In short, when comparing DOC patients to HCs, our findings reveal a significant decrease in CBF and metabolism, both globally and in seven networks. Moreover, machine learning analyses demonstrated that classifiers trained on either ASL or on FDG-PET data exhibited comparable diagnostic performance in distinguishing between DOC patients and HCs. These findings suggest a high degree of similarity between ASL and FDG-PET in diagnostic accuracy. When considering the relative advantages of ASL over FDG-PET, we propose that ASL may be a viable alternative to FDG-PET for clinical diagnosis of DOC. The results also highlight the potential utility of this method in the context of other neurological disorders.

## Data Availability

All data produced in the present study are available upon reasonable request to the authors

## Acknowledgments

The authors thank all the participants, along with the families or legal guardians of the DOC patients. We also thank the support staff at TMU Shuang-Ho Hospital for their unfailing assistance, in particular the Departments of Neurology, Neurosurgery, Radiology, Nuclear Medicine, and Rehabilitation. This work was funded by grants to TJL from the Taiwan National Science and Technology Council (105-2632-H-038-001-MY3, 106-2410-H-038-003-MY3, 111-2410-H-038-012), as well as by donations to the Brain and Consciousness Research Centre (Taiwan Ministry of Health and Welfare registration number: T063). NWD acknowledges support from the Taiwan National Science and Technology Council (110-2628-H-038-001-MY4).

## Author Contributions

Conception and Design: TJL. Data Analysis: JH, NWD. Data Collection: TJL, LH, CMY, CL, YCW, DY-TC. Data Curation: CL, NWD. Clinical Care: LH. Manuscript Composition: JH, TJL, NWD. Patient Assessment: LH, TJL, CL. Logistics and supervision: TJL. All authors reviewed and approved the final manuscript.

## GLOSSARY

ASL: arterial spin labeling
AUC: area under the curve
CBF: cerebral blood flow
CRS-R: Coma Recovery Scale-Revised
CSF: cerebrospinal fluid
DOC: disorders of consciousness
DAN: dorsal attention network
DMN: default mode network
FDG-PET: [^18^F]-fluorodeoxyglucose-positron emission tomography
FPN: frontoparietal network
LN: limbic network
MCS: minimally consciousness state
pCASL: Pseudo-Continuous Arterial Spin Labeling
ROC: receiver operating characteristic
ROIs: regions of interest
SMN: somatomotor network
SVM: support vector machine
UWS: unresponsive wakefulness syndrome
VAN: ventral attention network
VN: visual network

